# Associations between COVID-19 transmission rates, park use, and landscape structure

**DOI:** 10.1101/2020.10.20.20215731

**Authors:** Thomas F. Johnson, Lisbeth A. Hordley, Matthew P. Greenwell, Luke C. Evans

## Abstract

The COVID-19 pandemic has had severe impacts on global public health. In England, social distancing measures and a nationwide lockdown were introduced to reduce the spread of the virus. Green space accessibility may have been particularly important during this lockdown, as it could have provided benefits for physical and mental wellbeing. However, the associations between public green space use and the rate of COVID-19 transmission are yet to be quantified, and as the size and accessibility of green spaces vary within England’s local authorities, the risks and benefits to the public of using green space may be context-dependent. To evaluate how green space affected COVID-19 transmission across 299 local authorities (small regions) in England, we calculated a daily case rate metric, based upon a seven-day moving average, for each day within the period June 1^st^ - November 30^th^ 2020 and assessed how baseline health and mobility variables influenced these rates. Next, looking at the residual case rates, we investigated how landscape structure (e.g. area and patchiness of green space) and park use influenced transmission. We first show that reducing mobility is associated with a decline in case rates, especially in areas with high population clustering. After accounting for known mechanisms behind transmission rates, we found that park use (showing a preference for park mobility) was associated with decreased residual case rates, especially when green space was low and contiguous (not patchy). Our results support that a reduction in overall mobility may be a good strategy for reducing case rates, endorsing the success of lockdown measures. However, if mobility is necessary, outdoor park use may be safer than other forms of mobility and associated activities (e.g. shopping or office-based working).

## 1. Introduction

The COVID-19 pandemic has had severe impacts on public health (Mahase, 2020) and remains an emergency of international concern. In response to the pandemic, the UK government implemented social distancing measures and nationwide lockdowns to control the spread of the virus (UK Government, 2020a). During these periods, the general public were limited in the distances they could travel and, at certain points, the number of times they could leave their residence each day; with an allowance of one non-essential trip during the peak of transmission (UK Government, 2020a). Though social restrictions have fluctuated in response to case rates, social distancing has been constant and there has been a general message of reduced movement and staying local where possible for much of 2020 and throughout 2021. These restrictions have meant that members of the public became more reliant on amenity spaces close to their residences for daily exercise and/or recreation (Geng et al., 2021). Green spaces may provide a comparatively safe place for these activities, though the amount and structure of green space available for public use differs widely across the UK. Here we evaluate if differences in the availability and structure of public green space within local authorities (local government bodies responsible for public services within a specified area) in England, and their usage, influenced the local rate of incidence of COVID-19.

Green spaces, which we define as vegetated non-arable areas - see Taylor & Hochuli (2017) for further details - provide important cultural and recreational ecosystem services, benefiting both mental and physical health (Beyer et al., 2014; Cohen-Cline et al., 2015). These benefits are usually considered in terms of reducing the prevalence or severity of conditions such as mental stress (Nutsford et al., 2013) or cardiovascular disease (Seo et al., 2019), and some of these benefits have continued throughout the pandemic (Slater et al., 2020; Soga et al., 2020). However, the influence of green space use on disease transmission rates has received less investigation, but is of great importance as green space use has increased rapidly during the pandemic (Venter et al., 2020). Furthermore, it is unclear how ‘safe’ green spaces are during periods of higher incidence especially in densely populated areas (Shoari et al., 2020).

We anticipate that green space could impact COVID-19 incidence in two ways: general health and wellbeing, and transmission. It is conceivable that general health and well-being is greater in areas with more green space, as higher levels of green space are associated with healthier populations (Maas et al., 2006; Mitchell and Popham, 2007; van den Berg et al., 2015). As COVID-19 has a greater impact on those with underlying health conditions and sedentary lifestyles (Hamer et al., 2020; Jordan et al., 2020), green space may, therefore, indirectly provide some level of resilience to the disease and/or reduce incidence. However, our key focus here is on transmission, as it is likely that the major benefits of outdoor recreation in green space are related to a lower risk of infection. Current evidence suggests that COVID-19 is spread via droplet infections, contact with contaminated individuals or surfaces, and through aerosol transmission (Bahl et al., 2020). These risks are likely minimised in green space areas, as generally, they are less spatially confined, and contain fewer surfaces prone to frequent touching or contact. Consequently, green space use may represent a safe form of recreation by minimising risk of infection.

In England approximately 87% of the population are within a 10-minute walk of public parks and gardens (Shoari *et al*. 2020). However, both the structure and amount of green space vary between local authorities, and both could influence COVID-19 incidence. Generally, it has been found that greater health benefits are derived from larger areas of green space (Ekkel and de Vries, 2017). In the context of disease transmission, larger areas may offer more space per individual, lowering transmission risk. However, smaller fragmented areas of green space are common in many residential areas and are, therefore, more accessible to much of the population and may be used more frequently. Further, if public use is distributed across fragmented green spaces, the wider effects of a transmission incident could be reduced, as contacts would be isolated to the members of a neighbourhood or community adjacent to a particular green space. This process can be seen in animal diseases where habitat fragmentation reduces transmission due to limiting interactions between groups in different patches (Mccallum and Dobson, 2002). However, fragmentation also typically results from reductions in the total area of green space (Fahrig, 2013), leading to less overall space per individual, possibly increasing transmission rates.

Whilst the effects of green space on COVID-19 transmission are currently unclear, other environmental and social factors are known to influence both the spread and severity of the disease. For example, human mobility drives the spread of infectious diseases (Kraemer et al., 2019) and studies have shown that reducing social interactions by restricting mobility can lead to a decrease in transmission rates of COVID-19 (Chinazzi et al., 2020; Gatto et al., 2020). Furthermore, as diseases are often spread along transport links and in offices (Gatto et al., 2020; Zhang et al., 2018), enforcing lockdown situations that curtail movement, such as requiring people to work from home, can have a great effect on reducing transmission rates. In addition to mobility, health and social factors have been associated with increased severity of the disease such as age, underlying health conditions, and deprivation (Richardson et al., 2020; Williamson et al., 2020). Consequently, any possible effects of green space must be considered after attempting to account for factors that could increase recorded incidence.

Given the stated benefits of green space, it is important to attempt to evaluate using the available evidence, the impact of green space use on transmission rates. In addition, understanding the influence of green space on COVID-19 incidence could provide an estimate of the value of green space for maintaining public health if subjected to a resurgence of the COVID-19 pandemic. And, in the longer term, indicate the potential benefits of local green space on future pandemics of comparative severity. Here, using time series of COVID-19 cases within local authorities in England, we explore how both green space use and access (i.e. availability of green spaces) influence COVID-19 incidence. Our approach is to first construct a baseline transmission model to attempt to control for factors likely to influence recorded COVID-19 incidence and then to explore how green space influenced case rates above or below this baseline. We predict that green space and the way it is structured will, in itself, have no effect on case rates. However, we expect that an increase in relative park use (i.e. spending time in green space over indoor activities) will make the structure and availability of green space important (Figure 1). Specifically, when green space is low, park use will likely represent a safer form of movement (e.g. compared to shopping), unless the green space becomes a congregation zone that inflates transmission risk. Furthermore, we predict that case rates will be lower when green space is fragmented, as the disease will be contained in more localised areas. For example, if the local authority has one large park the presence of an infected individual puts more people at risk than an infected individual attending one of many parks. Further, we predict, as others have found (Kraemer et al., 2020), that increased mobility will increase incidence, but that park use (measured as relative use of parks) is a relatively safe form of mobility (e.g. preferable over shopping).

**Figure 1.**
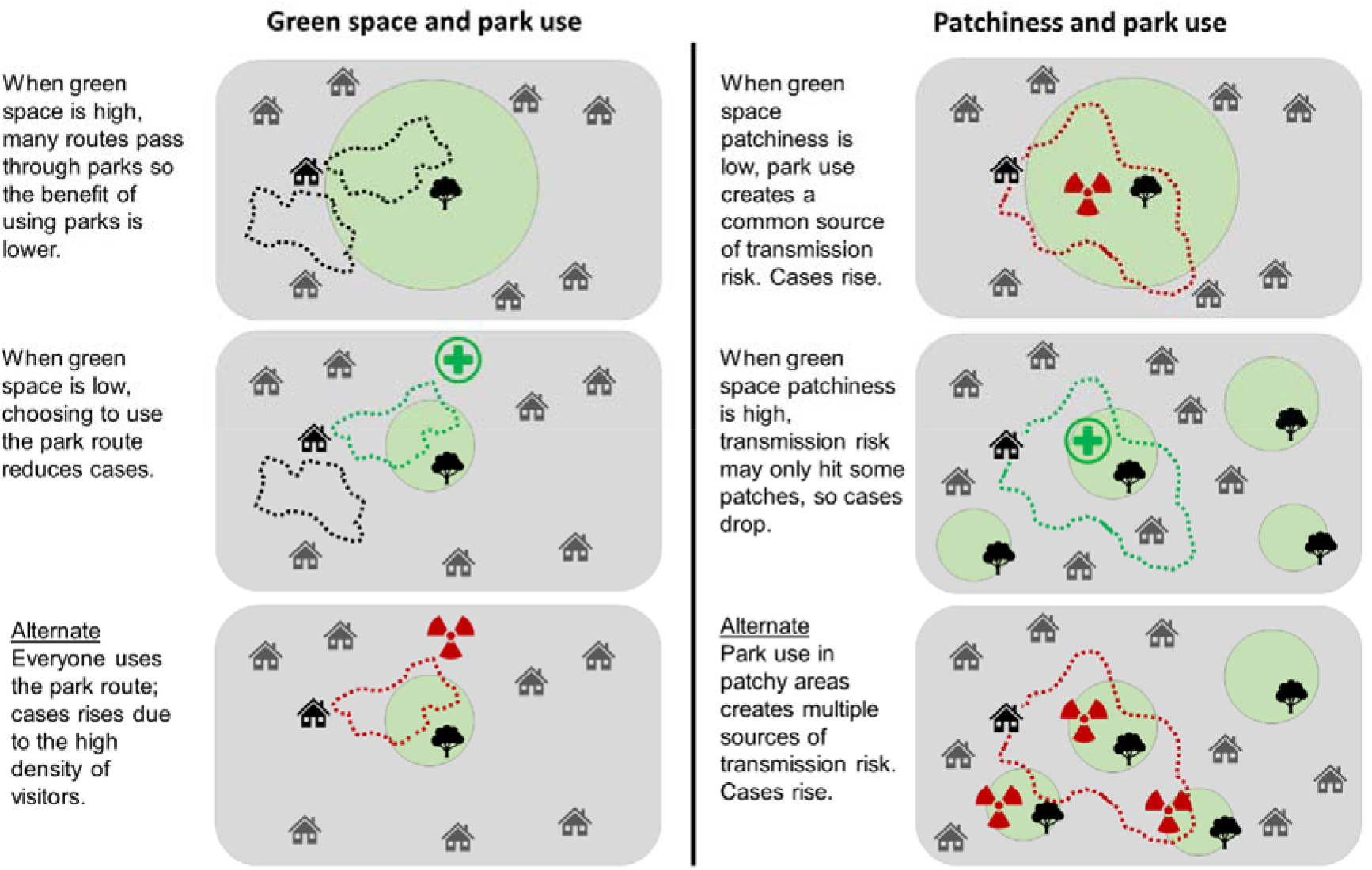
Mechanisms by which green space and patchiness could interact with park use to influence COVID-19 transmission. The upper two rows describe the primary predictions, whilst the bottom row explains alternate predictions. All variables (e.g. population density) except green space and patchiness, respectively, are held at a constant in these predictions. Green circles with a tree icon indicate the presence of green space. Dotted lines indicate walking routes, which becomes park use when the line overlaps a green space. The green health symbol indicates that the landscape metric and park use is beneficial, whilst the red toxic symbol indicates a risk.

## 2. Methods

### 2.1 Data compilation

#### 2.1.1 COVID-19 case rates

We compiled daily lab-confirmed cases (incidence) of COVID-19 in England from February 15^th^ 2020 up to November 30^th^ 2020 (available from https://coronavirus.data.gov.uk/). We only included cases until November, as in December England began an aggressive vaccination campaign and the more infectious COVID B1.1.7 variant began to spread widely (Horby et al., 2021) – factors that could confound our models (see below). Cases were recorded at the lower tier local authority (administrative areas for local government) level (N = 299). These local authorities vary in size (3 – 26,000km^2^), demographics, cultures, and in socio-economic circumstances. Incidence over this time was highly variable with periods of rapid increases, which were then relatively controlled by periods of national lockdown (Figure 2). To determine factors influencing COVID-19 transmission, we estimated case rates for each day in each local authority. Case rates were derived by fitting log-linear models, regressing the natural log of daily cases against date (days). To reduce the effect of daily variation in reported cases and instead capture the general trend, we fit these regressions over 7-day moving windows (Figure S1) e.g. to estimate the case rate on August 4^th^, a regression was fit between cases from August 1^st^ – 7^th^, for August 5^th^ a regression was fit between August 2^nd^ – 8^th^. The coefficients of these models provided a daily case rate. We converted these coefficients into a daily percentage change in cases. We opted to calculate case rates instead of using raw daily case numbers, as case rates more adequately capture transmissibility i.e. regardless of whether cases jumped from 5 to 10, or 50 to 100, the case rates would capture the doubling effect. Furthermore, case rates are more robust to variation in the population size of a local authority.

**Figure 2.**
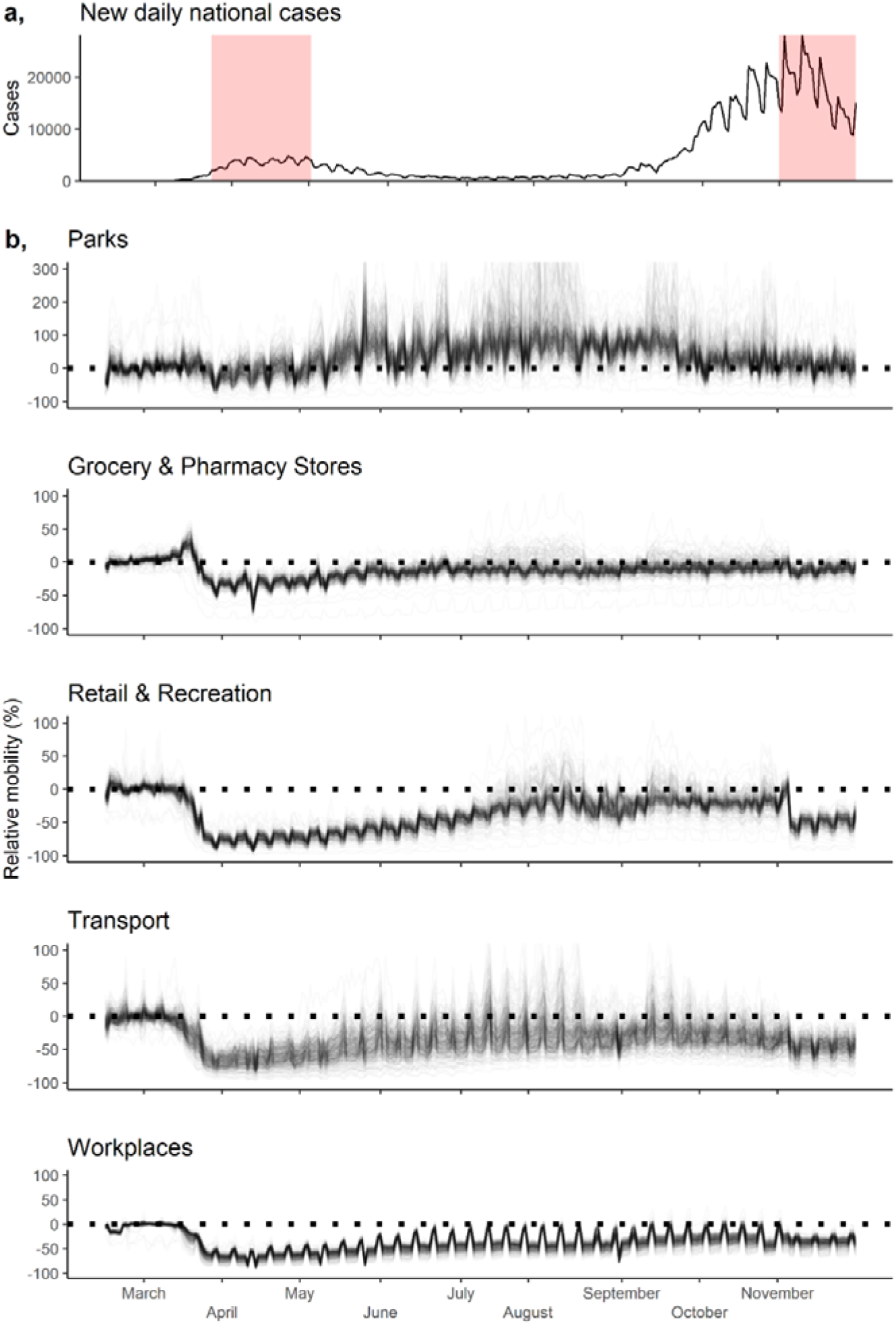
a) Daily lab-confirmed cases across England, with lockdown periods (with restricted mobility) indicated with red shading. b) Google mobility trends (Google, 2020), describing change in mobility over time for five different categories, relative to a baseline period (January 3^rd^ to February 6^th^ 2020). We excluded the sixth category ‘residential mobility’ as it is measured differently to all other categories (Google, 2020). Each line within the mobility trends represents a local authority. All plots extend from February 15th to November 30^th^ 2020. For the ‘parks’ plot, we limited the y-axis at 300% to exclude a small number of extreme observations with high park use.

#### 2.1.2 Baseline transmission variables

We compiled variables which describe the mechanisms considered to influence case rates (Table 1). Firstly, we derived two variables which describe the structure of the local authority population: population density – residential population density (controls for green space in the green transmission model below); and population clustering – Moran’s I spatial autocorrelation of residential population density (controls for patchiness in the green transmission model below). Secondly, we compiled three variables which characterise the human population in each local-authority prior to COVID-19: health – risk of premature death or a reduction in quality of life due to poor mental or physical health (Ministry of Housing Communities & Local Government, 2019); demography - the percentage of the population over 70 (Office for National Statistics, 2021a); economy – the percentage of unemployed-individuals in the non-retired local authority population (UK Government, 2018). A high baseline health, whereby few individuals have pre-existing underlying health conditions, may decrease the chances of an individual presenting with severe symptoms of COVID-19 and further passing the virus to others (Clark et al., 2020). Accounting for this baseline health may also assist in controlling for the presence of asymptomatic undetected infections in case rates.

**Table 1.**
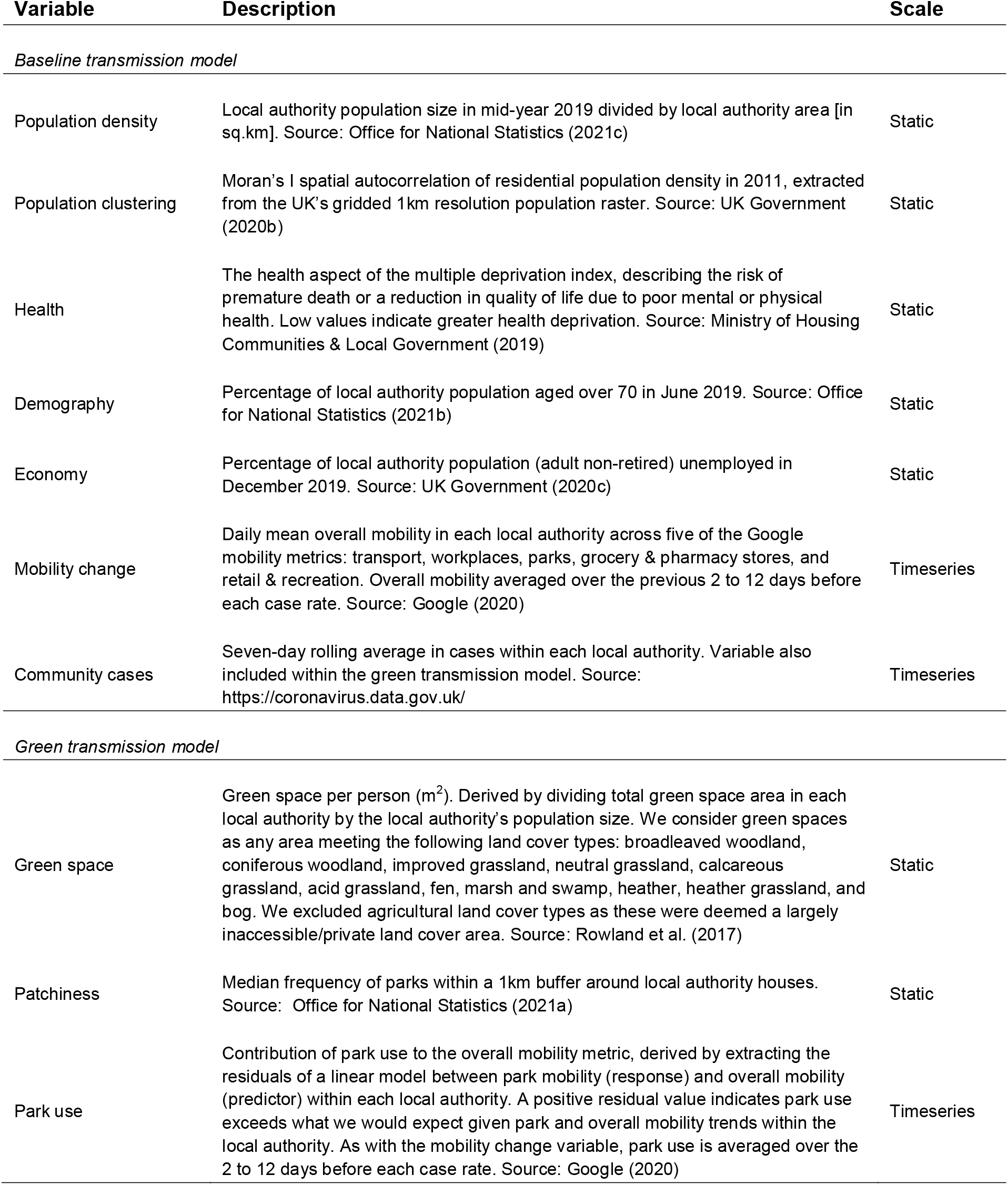
Description of variables within the baseline and green transmission models, including the scale at which the variable is measured, where ‘Static’ indicates only one value is derived per local authority, whilst there are unique values for each case rate in ‘Timeseries’ variables.

| Variable | Description | Scale |
| --- | --- | --- |
| *Baseline transmission model* |  |  |
| Population density | Local authority population size in mid-year 2019 divided by local authority area [in sq.km]. Source: Office for National Statistics (2021c) | Static |
| Population clustering | Moran's I spatial autocorrelation of residential population density in 2011, extracted from the UK's gridded 1km resolution population raster. Source: UK Government (2020b) | Static |
| Health | The health aspect of the multiple deprivation index, describing the risk of premature death or a reduction in quality of life due to poor mental or physical health. Low values indicate greater health deprivation. Source: Ministry of Housing Communities & Local Government (2019) | Static |
| Demography | Percentage of local authority population aged over 70 in June 2019. Source: Office for National Statistics (2021b) | Static |
| Economy | Percentage of local authority population (adult non-retired) unemployed in December 2019. Source: UK Government (2020c) | Static |
| Mobility change | Daily mean overall mobility in each local authority across five of the Google mobility metrics: transport, workplaces, parks, grocery & pharmacy stores, and retail & recreation. Overall mobility averaged over the previous 2 to 12 days before each case rate. Source: Google (2020) | Timeseries |
| Community cases | Seven-day rolling average in cases within each local authority. Variable also included within the green transmission model. Source: https://coronavirus.data.gov.uk/ | Timeseries |
| *Green transmission model* |  |  |
| Green space | Green space per person (m<sup>2</sup>). Derived by dividing total green space area in each local authority by the local authority's population size. We consider green spaces as any area meeting the following land cover types: broadleaved woodland, coniferous woodland, improved grassland, neutral grassland, calcareous grassland, acid grassland, fen, marsh and swamp, heather, heather grassland, and bog. We excluded agricultural land cover types as these were deemed a largely inaccessible/private land cover area. Source: Rowland et al. (2017) | Static |
| Patchiness | Median frequency of parks within a 1km buffer around local authority houses. Source: Office for National Statistics (2021a) | Static |
| Park use | Contribution of park use to the overall mobility metric, derived by extracting the residuals of a linear model between park mobility (response) and overall mobility (predictor) within each local authority. A positive residual value indicates park use exceeds what we would expect given park and overall mobility trends within the local authority. As with the mobility change variable, park use is averaged over the 2 to 12 days before each case rate. Source: Google (2020) | Timeseries |

National lockdowns, and the resulting reduction in people’s mobility, were an important tool for reducing transmission within England during the COVID-19 pandemic. We used Google Community Mobility Reports to track human mobility and its effect on case rates (Google, 2020). These reports chart movement trends over time across six categories: retail and recreation, groceries and pharmacies, transit stations, workplaces, residential, and parks. These trends describe how visitors to, or time spent in, each of the six categories changed compared to a pre-pandemic 5-week period (the median value from January 3^rd^ to February 6^th^ 2020). As the mobility data contained missing values (c.12%) for some local authorities and dates (Figure S2), we were conscious that these missing values may lead to statistical inference errors within the models below. As a result, we filled missing mobility values using *mice: multiple imputation chained* equations R package and ‘2l.pan’ imputation approach, which is a hierarchical normal model within homogenous within group variances (Van Buuren and Groothuis-Oudshoorn, 2011). This hierarchical structure allowed us to model mobility trends accounting for differences in local authorities. We included the following terms within our imputation model: five Google mobility timeseries (all except residential), as well as a 1-day lag period for each timeseries, the number of days along the timeseries since February 15^th^ with a cubic polynomial term, an indicator variable to describe whether each day was a weekend or not, and the timeseries of daily COVID-19 cases within the local authority. We also included terms that didn’t vary through time, including: the latitude and longitude of the local authority, and all local authority covairates within the baseline and green transmission models below (population density, population clustering, health, demography, economy, green space, and patchiness). Finally, we also included some national metrics that could infleunce local mobility, including: a timeseries of daily COVID-19 cases measured at the national scale, as well as the mean daily temperature and precipitation within Central England. We ran this model through 10 chains, each with 20 iterations, and 20 pan iterations. The imputation model converged.

Conventionally, as part of a multiple imputation framework, these 10 chains should then be modelled seperately and coefficient standard errors should be inflated with Rubin’s rules (Little and Rubin, 2002). However, given the small percentage of missing values, and that there are currently no well defined steps for using Rubin’s rules in genralized additive models (see our models below), we instead averaged mobility values across the 10 chains to produce mean estimates of mobility for each category, day, and local authroity i.e. conducting single impuation. We ensured the imputations produced plausible values (Figure S3). From this mobility dataset, we derived a variable which described overall mobility change for each date in each local authority, which is the average mobility change across five of the six categories (excluding residential) for each day in each local authority. We excluded the residential mobility category as it is inversely correlated with all other categories and is measured differently (Google, 2020). However, as there is likely a delay between a mobility reduction and a case rate reduction (Lauer et al., 2020), we lagged the overall mobility change metric by linking each case rate with the mean mobility change from 2 – 12 days prior. As a result of this lag, we trimmed the temporal extent of dataset to cover March 1^st^ – November 30^th^ 2020 (instead of February 15^th^ – November 30^th^ 2020).

#### 2.1.3 Green variables

We compiled two variables which describe the structure of green spaces in each local authority: patchiness – median frequency of parks within a 1km^2^ radius around households in the local authority (Office for National Statistics, 2021c); green space – available green space per person (m^2^) within the local authority, derived by dividing the green-cover area by the local authority population size. Green-cover area was calculated from the UKCEH 2015 25 metre land cover raster (Rowland et al., 2017) and covered a variety of landscape categories (Table 1). For this green-cover area calculation, we set a 1km buffer around the local authority, to represent green space access of households on the local authority border.

Using the mobility dataset, we also produced a park use variable, which represents how parks are used relative to overall mobility. This park use metric is derived by fitting a linear model between park use and overall mobility within each local authority, and extracting the residual park use, where positive values represent a preference for using parks over other forms of mobility for a given date (in the original percentage units). Parks include public gardens, castles, national forests, campsites, observation points, and national parks, but exclude surrounding countryside in rural areas. As a result, the Google (2020) definition of parks differs slightly to the landscape categories used in our green space metric but was our best available representation of how green space was used during the pandemic. As in the overall mobility change metric, park use represents the mean use of parks in the prior 2 to 12 days.

### 2.2 Modelling

We developed two core models (Figure 3): baseline transmission – aimed at controlling for the major mechanisms influencing case rates; and green transmission – impact of landscape structure and park use on case rates. The baseline and green transmission models are both focussed on case rates, but we anticipated that any effects of green space on COVID-19 case rates were likely to be much smaller than variables known to influence disease transmission (e.g. population density). As a result, we structured our analyses to first account for the presence of these more influential variables in a baseline transmission model, and then in the green transmission model, we explored how green areas (the focus of this study) can alter the residuals of these case rates. Conventionally, it is advised to include all variables within one regression instead of analysing the residuals separately (Freckleton, 2002). However, variables were highly correlated (e.g. population density and green space are derived in similar ways), and resulted in multicollinearity issues. Dealing with the major mechanisms first (e.g. population density) mitigated these multicollinearity issues.

**Figure 3.**
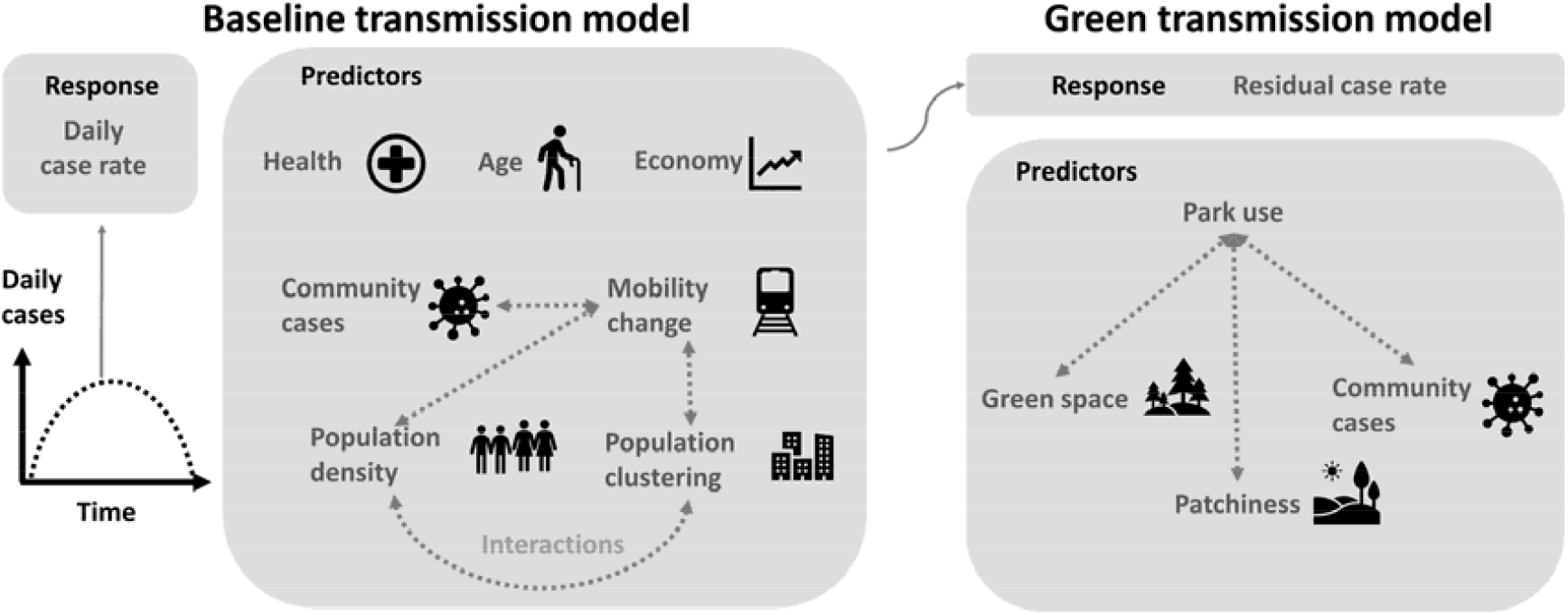
Model structure for baseline transmission and green transmission difference models, depicting the process for developing the response variables, as well as the predictors used in each model.

To control for the baseline health and transmission mechanisms influencing COVID-19 case rates, we developed a generalized additive model within the *mgcv* R package (Wood, 2021), with case rate as the response – inverse hyperbolic sine transformed to address heavy tailed residuals. We included the following parameters as linear predictors: health, demography, economy, population density (log_10_ transformed), population clustering, and mobility change. We also included interactions between population density and clustering, population density and mobility change, and population clustering and mobility change. In model development, it was clear that the residuals were experiencing extreme positive temporal autocorrelation, where case rate values were very similar to values from the previous day. As a result, we also included the previous days case rate (one day lag) as a linear predictor in the model. We included random intercept smoothing over the local authorities to account for the non-independence of multiple case rates within the same local authorities. Due to working hour restrictions in England, case counts on Saturdays and Sundays were largely underestimated, and then over-estimated on Mondays and Tuesdays. As a result, we also included a cyclic smoothing term (with up to 7 knots) over day of the week to capture reporting biases and control for daily variation (days within a week) in case reporting. We extracted the residuals from this model for the green transmission model.

To assess how landscape structure and park use influenced residual case rates, we again developed a generalized additive model, with residual case rates form the baseline transmission models as the response, as well as the following linear predictor parameters: park use, green space (log_10_ transformed), patchiness, as well as interactions between park use and green space, and park use and patchiness. These models also included random intercept smoothing over local authorities, but we did not control for the smoothing over days of the week, which was addressed in the earlier baseline transmission model.

#### 2.2.1 Sensitivity analysis

In both the baseline and green transmission models, we were conscious that some parameter effects may have varied through time. For example, some covariates may have been particularly influential prior to mandatory mask wearing in shops on July 24^th^ 2020. As a result, we extracted the first four weeks of data from our case rate dataset and ran the models on this subset. We then shifted the data forwards one week and re-ran the models, repeating this procedure (moving window), creating 40 replicates of the coefficients each representing a different-overlapping period of time between March 1^st^ and November 30^th^ 2020. From this, we established that the majority of coefficients were very stable over time (Figure S4), but mobility change, health, case rate lag, and park-use were somewhat variable. Looking at how these coefficients change through time, it was clear that mobility change had a temporal trend, where mobility effects were greatest when cases were at their highest. As a result, we amended the baseline transmission model to include an interaction between the mobility variables and the number of cases (averaged over the nearest 7 days) in the local authority at a given moment in time (see Equation S1-2 for the final model structures). There was no clear temporal trend in the health, case rate lag, and park-use variables so these remained untouched within the models. We also noted that the magnitude of the mobility change effect was far greater in the first lockdown period (March – May 2020). We suspect the large effect is genuine, but given there were spatial biases in case-testing availability during the first lockdown, we opted to re-model the data with a trimmed temporal extent (June 1^st^ to November 30^th^ 2020). From this, it was apparent that coefficients were generally far more conservative using the trimmed dataset, albeit still in the same direction (Figure S5). Given this discrepancy in results (depending on the temporal extent), we opted to restrict our analyses throughout the rest of this manuscript to solely focus on the more conservative trimmed temporal extent, which is likely to be far less effected by spatial variability in case-testing availability – so more robust. As a result, all model outputs and projections (see below) are derived from the data covering June 1^st^ to November 30^th^ 2020.

In the analyses, we opted to fill missing mobility values with imputation instead of using complete-case analyses, where any observations with missing mobility data are removed. However, given the small percentage of missing values, and that the mobility data is averaged across five categories, and then again through time, we wanted to ensure model coefficients did not change drastically under imputation, which could be a sign of a statistical inference error (Johnson et al., 2021). As a result, we repeated the analyses using only complete-case observations and compared model coefficients between the missing value approaches. Given the similarity in the complete-case and imputation coefficients (Figure S5), we continued using the coefficients from the imputation model which covered a greater array of local authorites.

#### 2.2.2 Model checking

We standardised (subtracting values from their mean and dividing by their standard deviation) all predictor variables in the models to determine effect sizes and reduce multicollinearity where interactions are present. All model assumptions passed e.g. multicollinearity (variance inflation factors less than 3 within both the baseline and green transmission model), concurvity (observed and estimated concurvity less than 0.1), absence of spatial (Moran’s I = 0.1) and temporal autocorrelation (Figure S6), homogeneity of variance, and normality of residuals. When summarising results, we report the mean ± standard deviation, and when describing model outputs we report the standardised slope coefficient and 95% confidence intervals. We also report the *R*^*2*^ for each model. All analyses were conducted in R 4.0.3 (R Development Core Team, 2020).

#### 2.2.3 Projecting cases

To understand how mobility patterns have influenced cases, we projected cases using the baseline and green transmission models under three scenarios: 1) cases under observed mobility patterns; 2) cases after a 20% reduction in each day’s overall mobility; 3) cases after a 20% increase in each day’s park use. We ran the baseline and green transmission models through each of the scenarios for every local authority between March 1^st^ and November 30^th^ 2020. We standardised all authorities so they had the same starting number of cases (10), community cases (10), and lagged case rate (0.58%; the mean case rate across local authorities on February 28^th^). These cases, community cases, and lagged case rate were updated and iteratively informed by the new model predictions, instead of the observed data. As a result, the projected case rates are solely influenced by the landscape structure and mobility patterns in the local authority. We constrained the case rates so they could not exceed the range of the observed case rates (−40% to 70%). We converted the projected case rates into projected cases, against the starting case value of 10.

## 3. Results

Across the 299 local authorities, case rates fluctuated substantially through time (Figure 4a). Mobility declined substantially during the first national lockdown in March to May, and in the run up to winter (Figure 4b). During the summer months, mobility and the variance in mobility increased, and in some local authorities these increases were close to 100% (doubling mobility). In contrast, park use increased during the first lockdown and remained high (approximately 25% above baseline) until winter approached in October (Figure 4c). There was less variation in park use trends between local authorities than in the mobility change metric.

**Figure 4.**
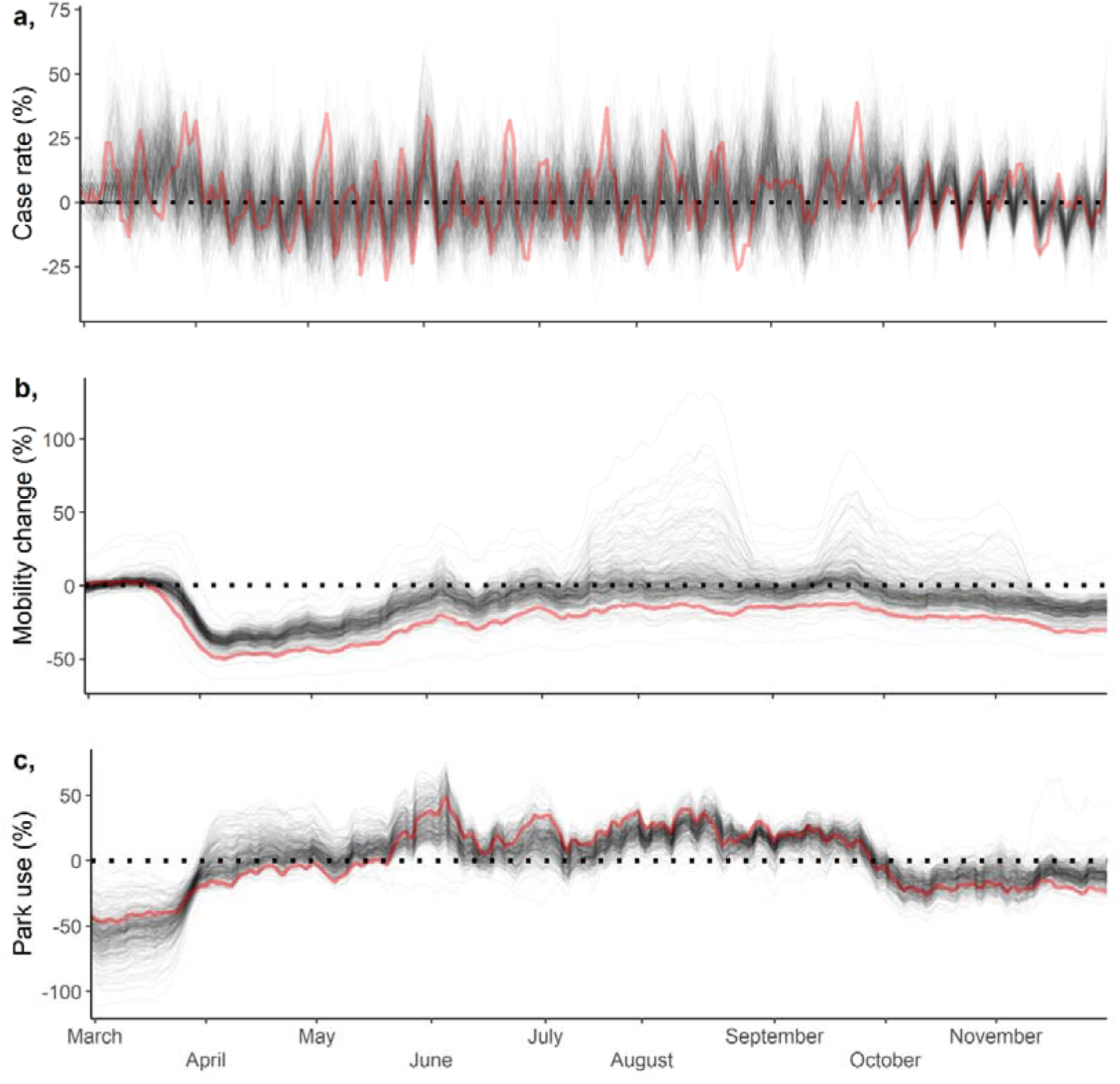
a) Temporal patterns in case rates (a), mobility change (b) and park use (c) between March 1^st^ and November 30^th^ 2020, with each line representing a different local authority. The red line represents the Oxford local authority and acts purely as an example. Case rates are defined as the daily percentage change in cases calculated over a seven day moving average. Mobility change is the mean daily percentage change over five mobility types (Park, Grocery and Pharmacy stores, Retail and recreation, Transport, and Workplaces) extracted from Google community mobility reports (Google, 2020). Park use is the relative contribution of park mobility to overall mobility change, derived by extracting the residuals of a linear model with park mobility regressed against overall mobility within each local authority i.e. are people visiting parks more than we would expect on a given date.

### 3.1 Baseline transmission models

Using the dataset with a trimmed temporal extent of June 1^st^ to November 30^th^ 2020 (see sensitivity analysis above), we observed an association between a reduction in mobility and a decline in case rates, and changes in mobility had a larger impact when there was a higher number of average cases and when the population was more clustered (Table 2; Figure 5c, d). Population density and population clustering had no significant impact on case rates. Increases in the health index and proportion of the population over the age of 70 were both associated with significant decreases in case rates (Table 2; Figure 5a, b). This baseline transmission model had an *R*^*2*^ of 0.45.

**Table 2.** Estimated regression parameters from the baseline and green transmission models with 95% confidence intervals. Values rounded to two significant figures, those with confidence intervals not overlapping zero (i.e. significant at the p = 0.05 threshold) are shown in bold. These coefficients were derived from models utilising the trimmed temporal extent dataset covering June 1^st^ to November 30^th^ 2020 – see sensitivity analysis above.

|  | Coefficient [95% confidence intervals] |
| --- | --- |
| *Baseline transmission model* |  |
| Intercept | **0.38 [0.36, 0.39]** |
| Lag case rate | **1.55 [1.53, 1.57]** |
| Population density | 0.020 [-0.006, 0.050] |
| Population clustering | 0.011 [-0.006, 0.028] |
| Mobility | **0.17 [0.15, 0.19]** |
| Case average | **0.061 [0.042, 0.080]** |
| Baseline health | **-0.031 [-0.054, -0.007]** |
| Percentage over 70 | **-0.051 [-0.079, -0.023]** |
| Percentage unemployed | 0.0027 [-0.024, 0.029] |
| Mobility:Case average | **0.11 [0.092, 0.13]** |
| Population density:Population clustering | 0.0060 [-0.011, 0.023] |
| Population density:Mobility | -0.011 [-0.025, 0.004] |
| Population clustering:Mobility | **0.029 [0.012, 0.047]** |
| *Green transmission model* |  |
| Intercept | 0.0001 [-0.016, 0.016] |
| Park use | **-0.057 [-0.074, -0.041]** |
| Green space | 0.0035 [-0.018, 0.025] |
| Patchiness | 0.010 [-0.011, 0.032] |
| Park use:Green space | **0.032 [0.010, 0.053]** |
| Park use:Patchiness | **0.024 [0.0026, 0.045]** |

**Figure 5.**
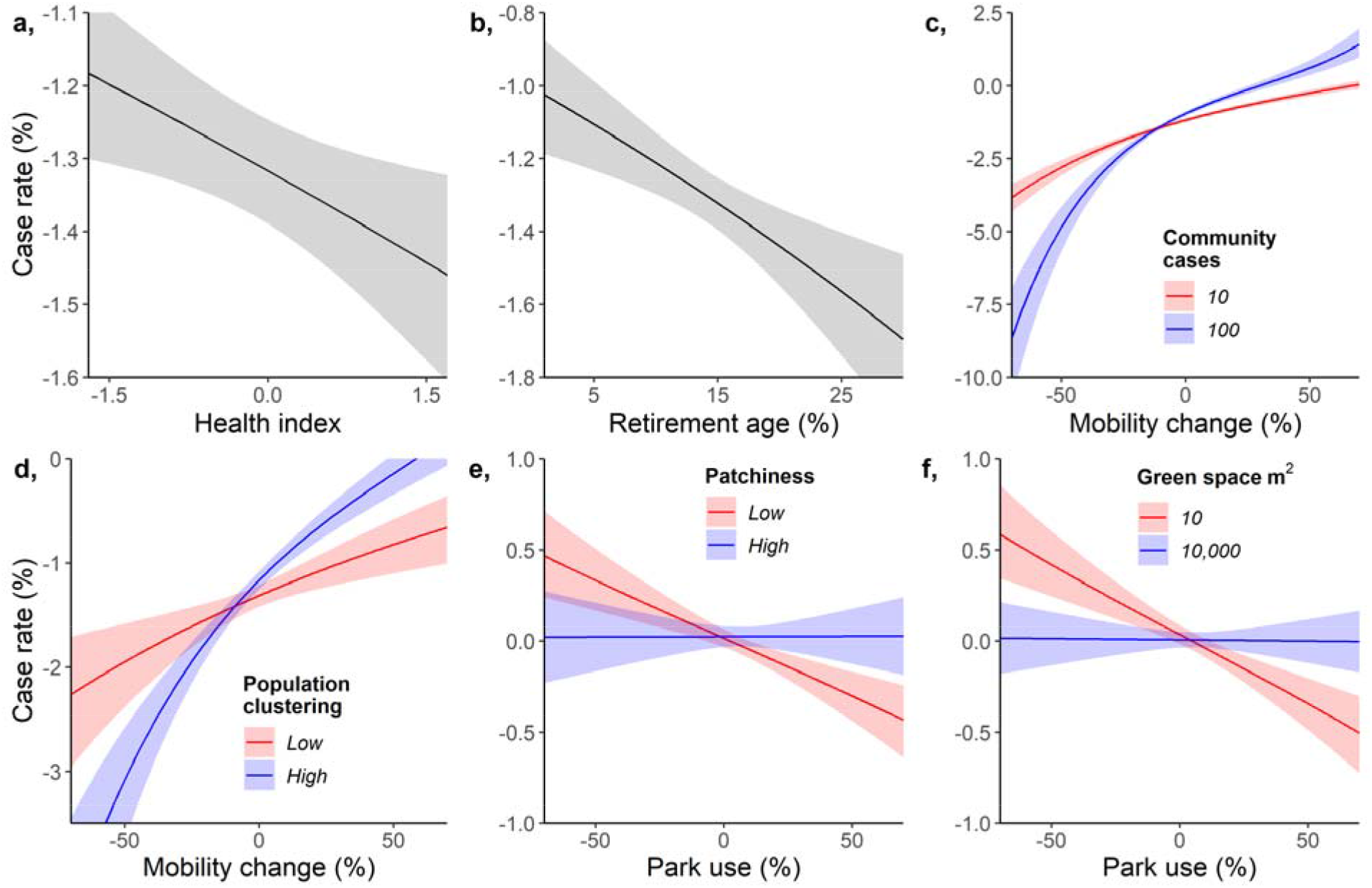
Marginal effects of important interaction parameters in the baseline transmission and in the green transmission models. Marginal effects are held at zero for all other parameters as variables were z-transformed. Panels depict the effect of: a) health, with low values indicating health deprivation; b) the percentage of the population over 70; c) an interaction between mobility and community cases (the 7-day average number of cases in the local authority); d) an interaction between mobility and human population clustering set at 0.2 (Low) and 0.7 (High), where 0 indicates a random distribution of clustering, and 1 indicates a complete separation in clustering; e) an interaction between park use and patchiness (the median frequency of parks within 1km of each house in a local authority); and f) an interaction between park use and green space area per local authority capita. Error bars represent the 95% confidence intervals. These marginal effect plots were derived from models utilising the trimmed temporal extent dataset covering June 1^st^ to November 30^th^ 2020 – see sensitivity analysis above

### 3.2 Green transmission models

Park use was associated with decreased residual case rates (Table 2; Figure 5e) but the size of the effect depended on the availability of green space and how patchy it was. When patchiness was high and when there was a large amount of greenspace, park use had less of an impact on case rates, though was still associated with a significant reduction in cases. The green transmission model had a small *R*^*2*^ of 0.01, despite the significant effects.

### 3.3 Projected cases

Reducing mobility is a far more effective measure of limiting COVID-19 transmission than increasing park use (Figure 6). Across local authorities between March 1^st^ and November 30^th^ 2020, a 20% reduction in mobility is projected to have led to 51% fewer cases on average (Figure 6b; 95% quantiles: -88.7% to -29.7%). In contrast, a 20% increase in park use is estimated to have only reduced cases by 5.4% (Figure 6c; 95% quantiles: -17.3% to 0.6%). So whilst park use is associated with reducing COVID-19 transmission, the benefits would only be relatively small. However, there is spatial variation in these findings, with some areas potentially benefitting more than others from a reduction in mobility or increase in park use (Figure 7).

**Figure 6.**
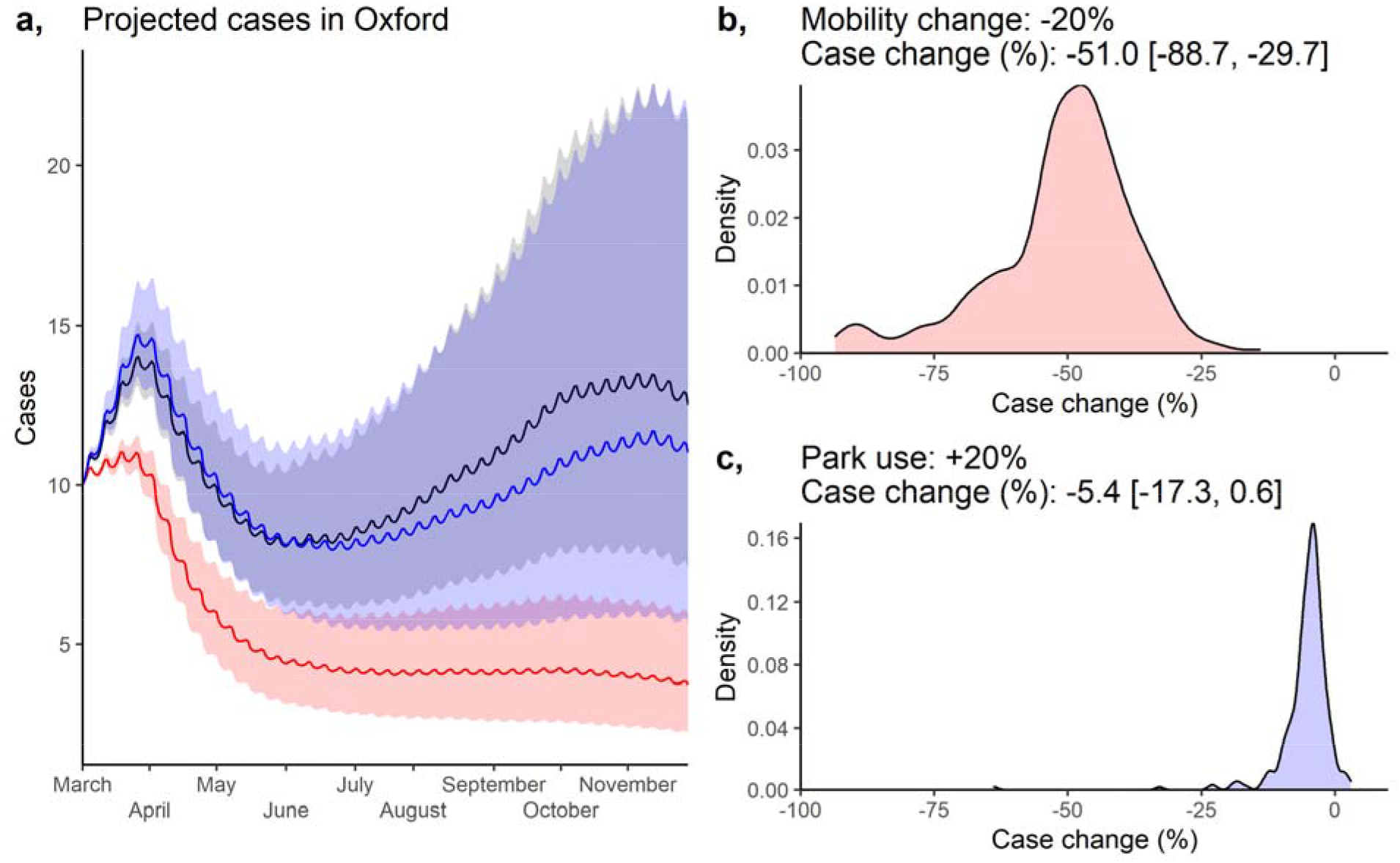
a) Projected daily cases between March 1^st^ and November 30^th^ 2020 within Oxford under three scenarios: 1) observed mobility patterns (black); 2) a further 20% reduction in observed mobility (red); and 3) 20% increase in observed park use (blue). In these projections, we set the initial cases (on March 1^st^) at 10, and with lagged case rate of 0.58% - the mean value across local authorities on February 28^th^. All other covariates were held at their observed values. Error ribbons represent 95% confidence intervals. Panels b and c represent the distribution of projected change in cases across local authorities under the 20% mobility reduction (b) and 20% park use increase (c) scenarios i.e. how much could cases have been reduced under these scenarios. Case change was derived by dividing the total cases between the March and November periods under each scenario by the cases in the observed mobility scenario (black), multiplying this value by 100, and then subtracting 100. Whilst these projections cover the period March 1^st^ – November 30^th^ 2020, the coefficients used to derive the projections were taken from the trimmed temporal extent dataset of June 1^st^ – November 30^th^ 2020, where coefficients were more conservative and less prone to bias (see *sensitivity analyses* above).

**Figure 7.**
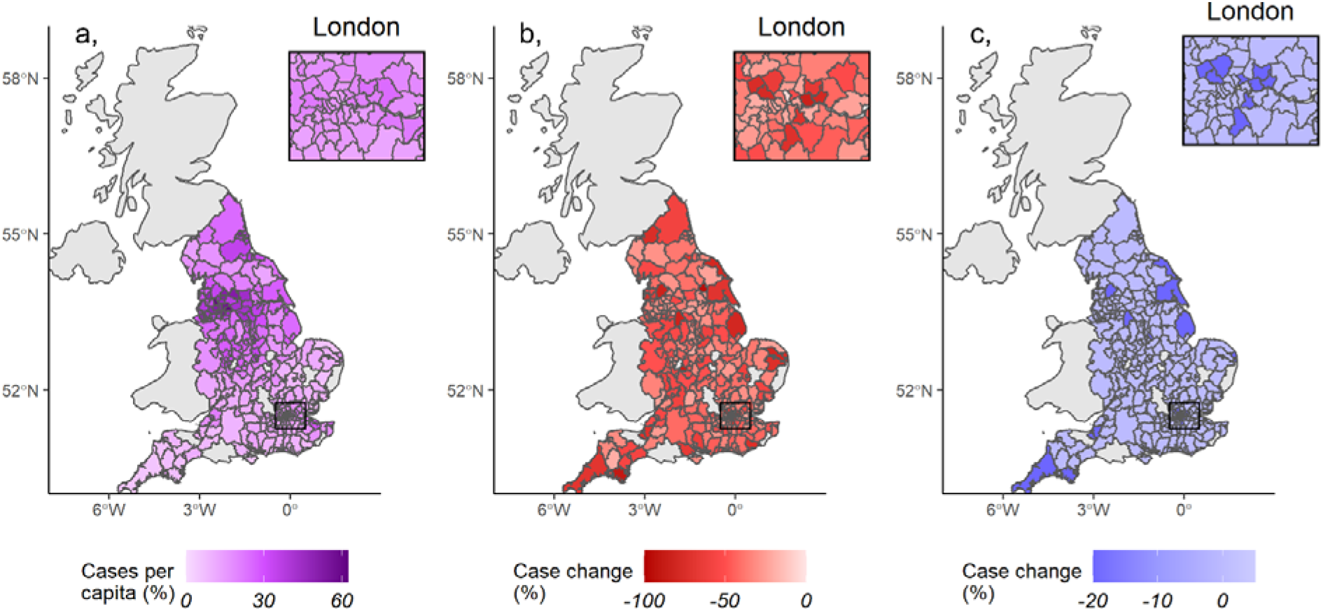
Spatial variation in observed cases per capita (a), and projected case changes under a 20% mobility reduction (b) and 20% increase in park use (c). Case change was derived by dividing the total cases between March and November 2020 under each scenario by the cases in the observed mobility projection, multiplying this value by 100, and then subtracting 100 (see Figure 6). The coefficients used to derive the projections in b and c were sourced from models utilising the trimmed temporal extent dataset covering June 1^st^ to November 30^th^ 2020 – see sensitivity analysis above

## 4. Discussion

In this study, we attempted to quantify the effects of local green space on COVID-19 case rates after accounting for mechanisms known to influence pandemics in our baseline transmission model. We found that high overall mobility was associated with increased case rates, especially when population clustering was high. After accounting for these variables, we found that higher park use, compared to other amenity areas, was associated with a reduction in case rates, especially in local authorities with low green space and with contiguous green space. These results suggest that utilising green spaces rather than carrying out other activities (e.g. visiting shops and workplaces) may reduce the transmission rate of COVID-19, but these benefits are limited compared to reducing mobility more generally.

From our baseline transmission model results, case rates were lower in local authorities with healthier populations and older populations (Figure 5a-b). These results are logical, firstly as previous evidence has shown COVID-19 has a greater impact on those with underlying health conditions (Hamer et al., 2020; Jordan et al., 2020) and more severe cases may be more likely to be tested and reported. Secondly, whilst the eldery are more at risk of mortality from COVID-19 (Williamson et al., 2020), this fact was widely reported in public health guidance and older people may have reduced contact with other individuals (Canning et al., 2020). Our baseline transmission model also shows that reducing mobility is most valuable when community cases are high and in areas with high population clustering (Figure 5c-d). This is consistent with person-person contact as the major mechanism of transmission and appears to demonstrate the general effectiveness of lockdown measures in reducing case rates, as others have demonstrated previously (Davies et al., 2020; Lau et al., 2020). Mobility had less impact in low clustered areas, which again may be expected, as people are more likely to be able to maintain distance and the potential number of interactions is reduced.

Once we had accounted for known drivers of case rates, we investigated how landscape structure and park use (i.e. mobility in green spaces) affected residual case rates using the green transmission model. Here we found that using parks, relative to other types of mobility, was associated with a reduction in case rates (Figure 5-6). However, reducing overall mobility (i.e. mobility to all amenity areas) led to a far more substantial decline in case rates. For example, a 20% reduction was projected to reduce cases by c.35%, whilst a 20% increase in park use was projected to reduce cases by 5% to 10% (Figure 6). This suggests that the use of parks may have modestly helped in reducing transmission rates in some areas during the pandemic, but reducing overall mobility is substantially more beneficial than maintaining mobility at pre-pandemic levels and spending that mobility in parks.

Whilst park use, overall, had a relatively small effect, we did note stronger effects of park use when the context of the local area was considered as using parks was beneficial in authorities with low green space and authorities with contiguous green space (Figure 5e-f and Figure 6). That park use has a minor beneficial effect overall seems to support our hypothesis that recreation in green space and parks may be safer than in other amenity areas. This is probably because it is easier to maintain distance and green spaces have fewer surfaces which could result in transmission if contaminated. However, the limiting impact of this when green space is high and accessible seems to suggest diminishing returns in how park use can impact COVID-19 transmission. This is perhaps not surprising if the main value of parks in this context is as an alternative to other relatively more hazardous amenity areas. Consequently, if there are other safe options outside of public parks then parks will likely have little impact. However, our findings do suggest that the use of public parks in a highly urbanised area may be advantageous, though as noted above the strongest effect was from the reduction of all forms of mobility. Therefore, cautiously, and given that it corresponds with common sense, we suggest that reducing mobility is a successful strategy for reducing case rates but given a need for some non-essential time outside of a home, using green spaces such as local parks may be the next best thing, particularly in highly urbanised areas.

A major limitation of the work is the difficulty in comparing across local authorities that vary simultaneously in many different variables likely important to case rates. This makes inference about the importance of their individual effects very difficult and so effect sizes should be interpreted cautiously and with caveat. Another challenge is that pandemics are rare events, consequently, our analysis covers only a snapshot of time for each local authority. During this period, many different factors not included in the analysis (e.g. chance super spreading events) may have explained much of the variation between local authorities. Despite this, the model fits are reasonably high. An additional limitation in our analyses is the absence of complete Google mobility data in some local authorities. We handled these missing values with imputation and attempted to ensure models were robust by comparing imputed models with complete-case models. Encouragingly, our complete-case and imputed results are very similar, which suggests the imputation has not introduced any missing data bias (Johnson et al., 2021) – although both the imputation and complete-case analysis could just be equally wrong. Given this uncertainty, and the further limitations we have identified above, our mobility findings should be interpreted cautiously.

One potential influence we failed to capture within our case rate modelling was the influence of environmental features like air pollution and weather. Air pollution has already been to linked to an increase in COVID-19 related deaths, and potentially even transmission (Travaglio et al., 2021). Similarly, there are plausible hypotheses that suggest weather effects like temperature, ultraviolet light, and wind speed may influence the virus’s persistence and in-turn transmission (Carlson et al., 2020). Importantly, both of these environmental features may also interact with the findings in our study. Firstly, park use may become a more inherently risky activity if air pollution at the green space is high. Secondly, as park use is likely very correlated with weather, the effects of park use may be confounded by weather. Both of these points warrant investigation, perhaps at a far finer scale than the local authority level.

Understanding the risks of different amenity areas could be important for longer-term management of COVID-19 and the landscape-dependency of this advice could be important for developing ‘local-lockdown’ guidance. In particular, access to green spaces has been shown to have benefits for mental and physical well-being (Slater et al., 2020; Soga et al., 2020), and consequently, understanding the relative risks of using these areas is important. Our results show that COVID-19 case rates may be reduced with individuals spending time in parks, relative to other amenity areas, especially in urbanised, high-density areas. Although further research is needed, these findings suggest that the use of parks for recreational activity in these contexts could be advisable, demonstrating a possible additional utility of these green spaces in addition to the known benefits to health and wellbeing (de Vries et al., 2003; Mitchell and Popham, 2007; Nutsford et al., 2013) in normal non-pandemic conditions.

## Data Availability

All data to repeat analyses are presented in the manuscript

## Acknowledgments

Thanks to the NERC Covid-19 hackathon for instigating this work. This work was partly funded by the following NERC (Natural Environment Research Council) Centre for Doctoral Training studentships: J71566E, P012345, and L002566.

## Data accessibility

Code and data to repeat analysis are presented in https://github.com/GitTFJ/COVID19_parks_landscape

## Author contributions

All authors contributed to project design. Analysis was led by TFJ and LCE, but all authors contributed. TFJ and LCE co-wrote the first draft and co-authors contributed to revisions.

