## Supplementary material for "Associations between COVID-19 transmission rates, park use, and landscape structure"

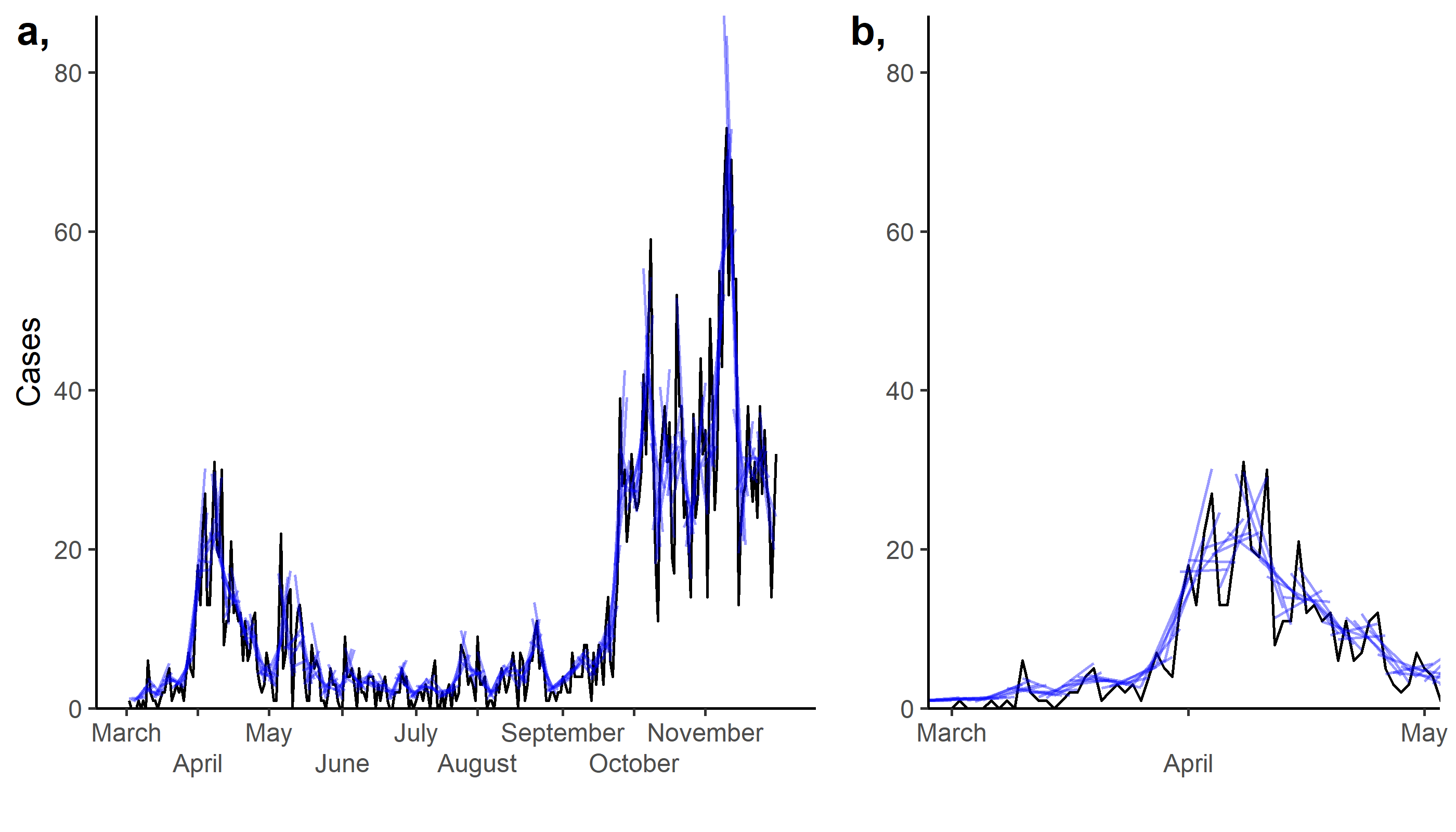

**Figure S1.** Approach for calculating case rates (example represents Oxford), with a seven-day moving window of linear models, regressing the natural log of cases against time (days since March 1^st^ 2020). Panel ‘a’ covers the full time frame whilst panel ‘b’ is a zoomed on version of the March to May period.

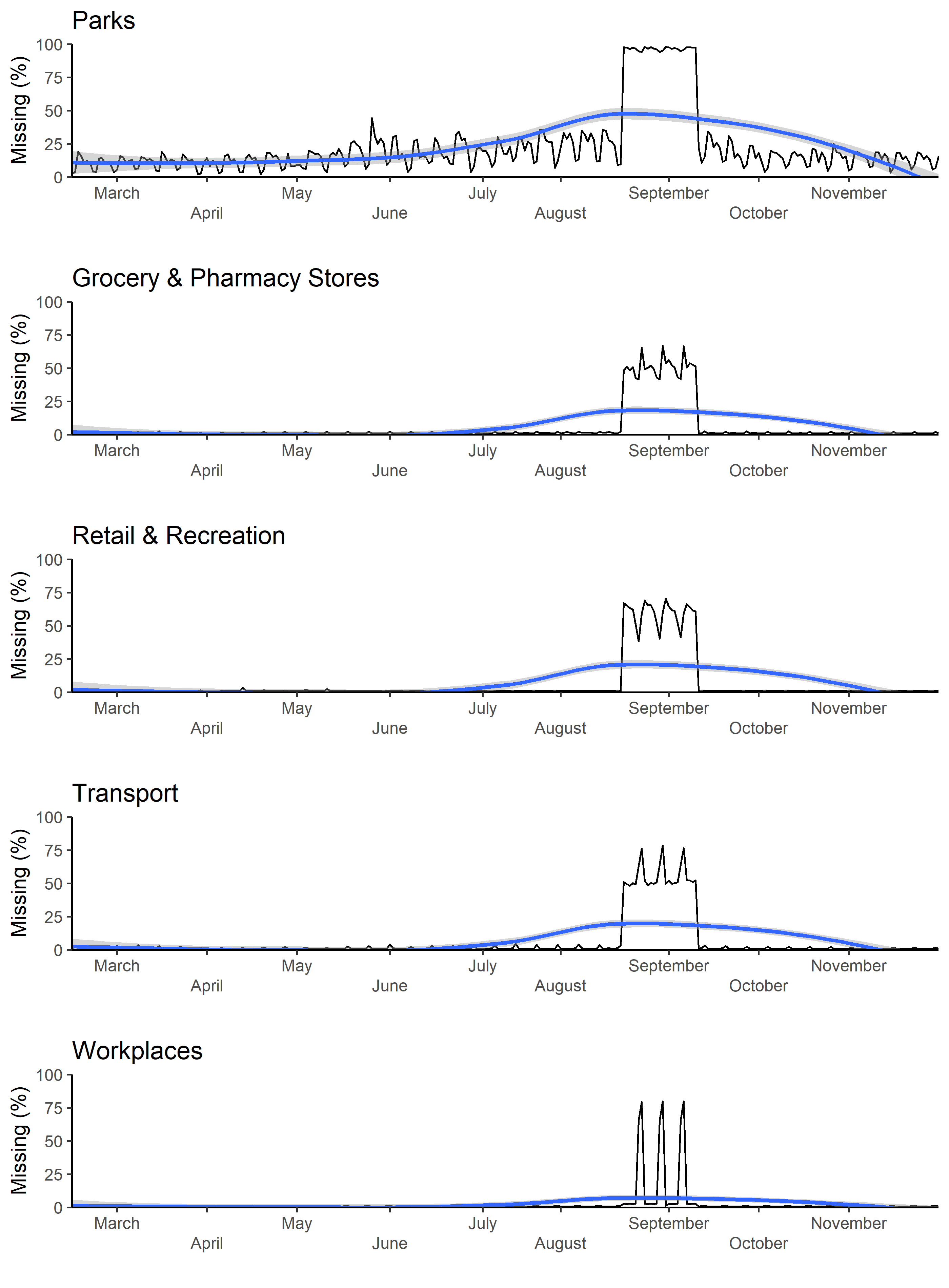

**Figure S2.** Percentage of local authorities missing mobility values in each day across the five mobility types used to calculate the mobility change (%) variable. The blue lines represent locally estimated scatterplot smoothing of the daily missing (%).

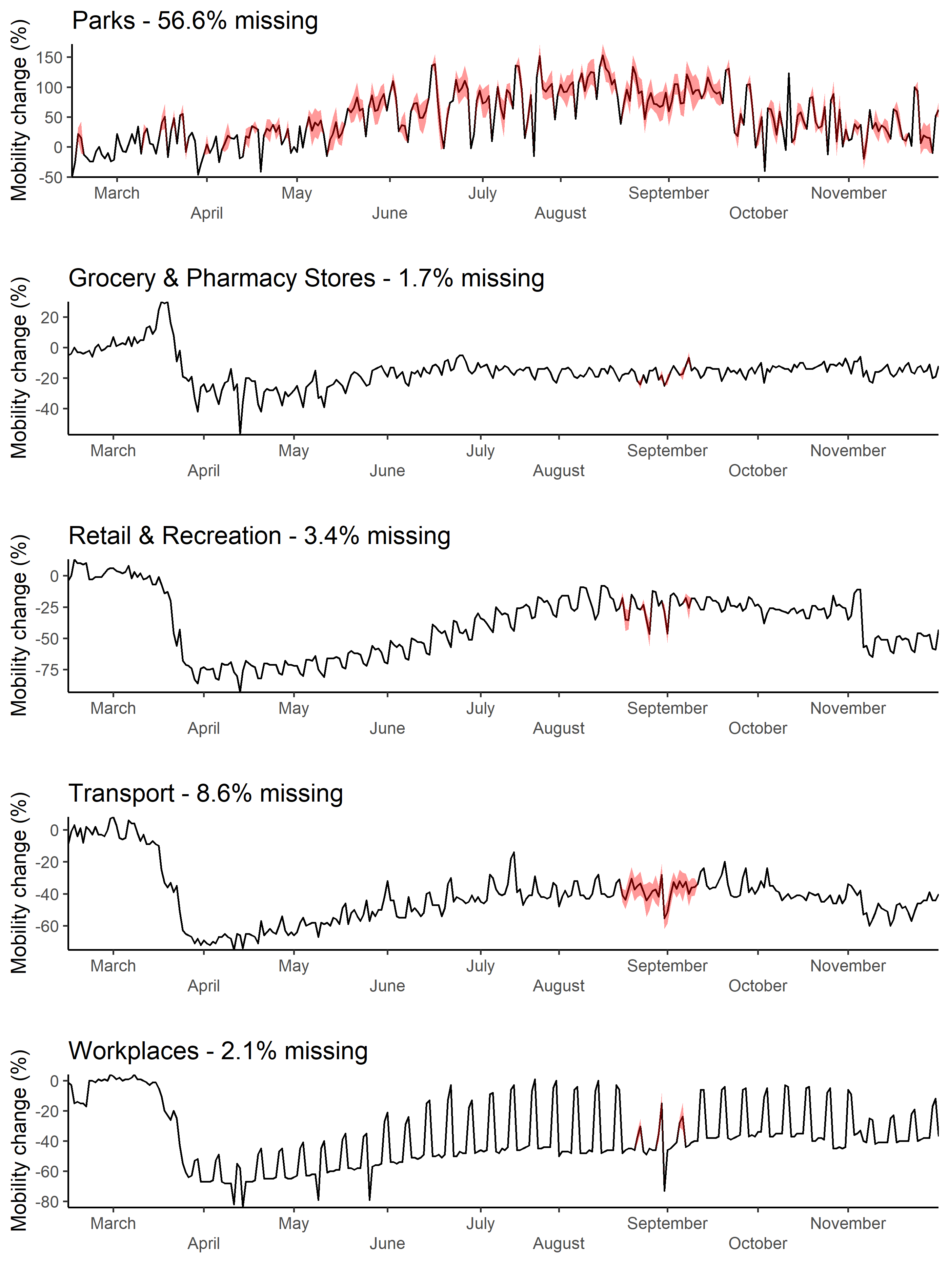

**Figure S3.** Estimated mobility for a local authority (Warrington) with a high proportion of missing mobility values. Mobility is split into the five categories used to derive the mobility change (%) variable. Missing values are indicated by red shading, which represents the 95% confidence intervals of each imputed value.

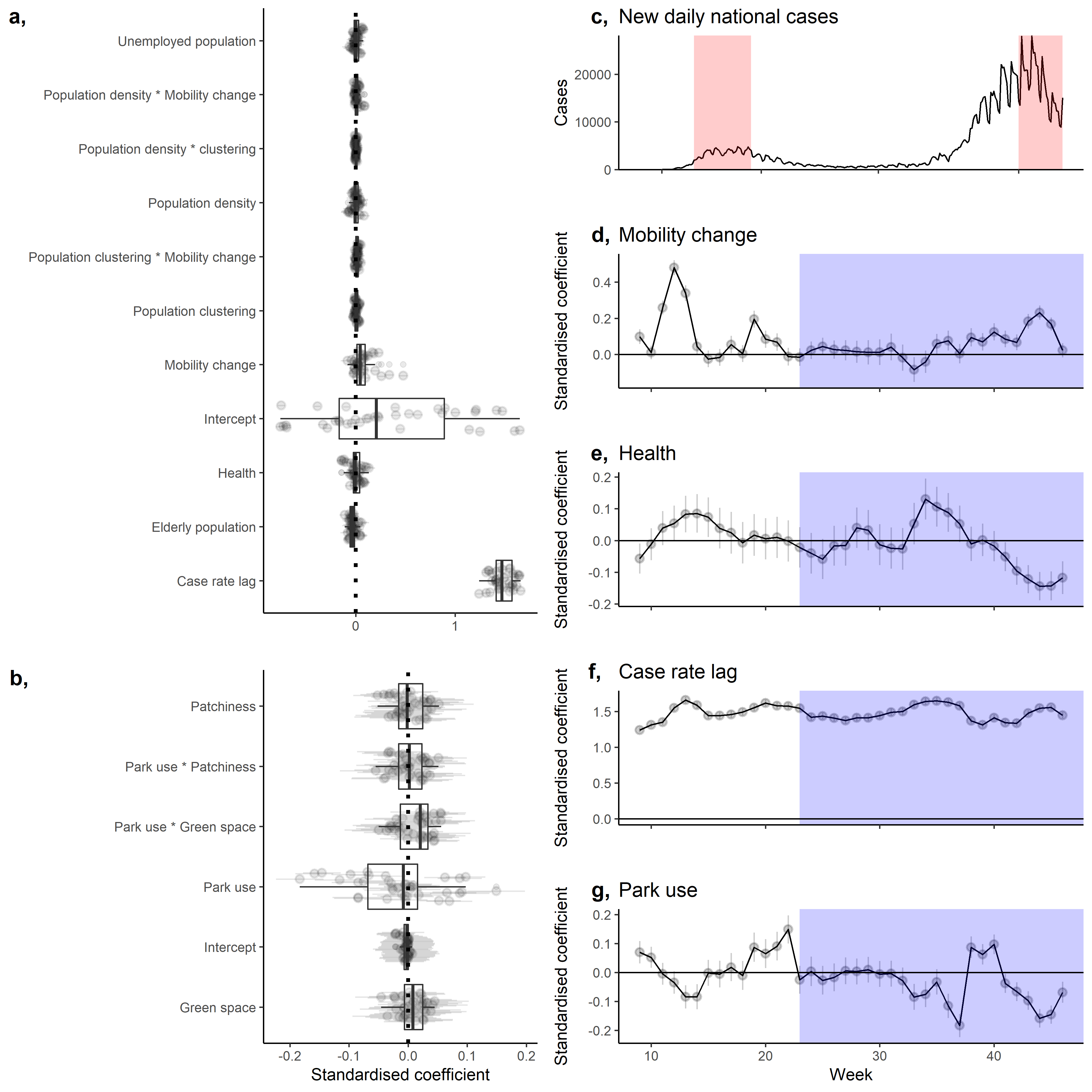

**Figure S4.** Standardised coefficients of the baseline (a) and green (b) transmission models. Each point represents a different four-week subsets of a moving window from the full dataset (March 1^st^ to November 30^th^ 2020; see sensitivity analysis in the main text for a description), with a boxplot summarising the median and quantiles of the coefficients. c) National daily lab confirmed cases in England with lockdown periods represented with red shading. Finally, temporal trend in mobility change (d), health (e), case rate lag (f), and park use (g) coefficients from the four-week moving window subsets. The blue shading in d-g represents the coefficients from just the trimmed dataset (June 1^st^ – November 30^th^ 2020; see sensitivity analysis in the main text).

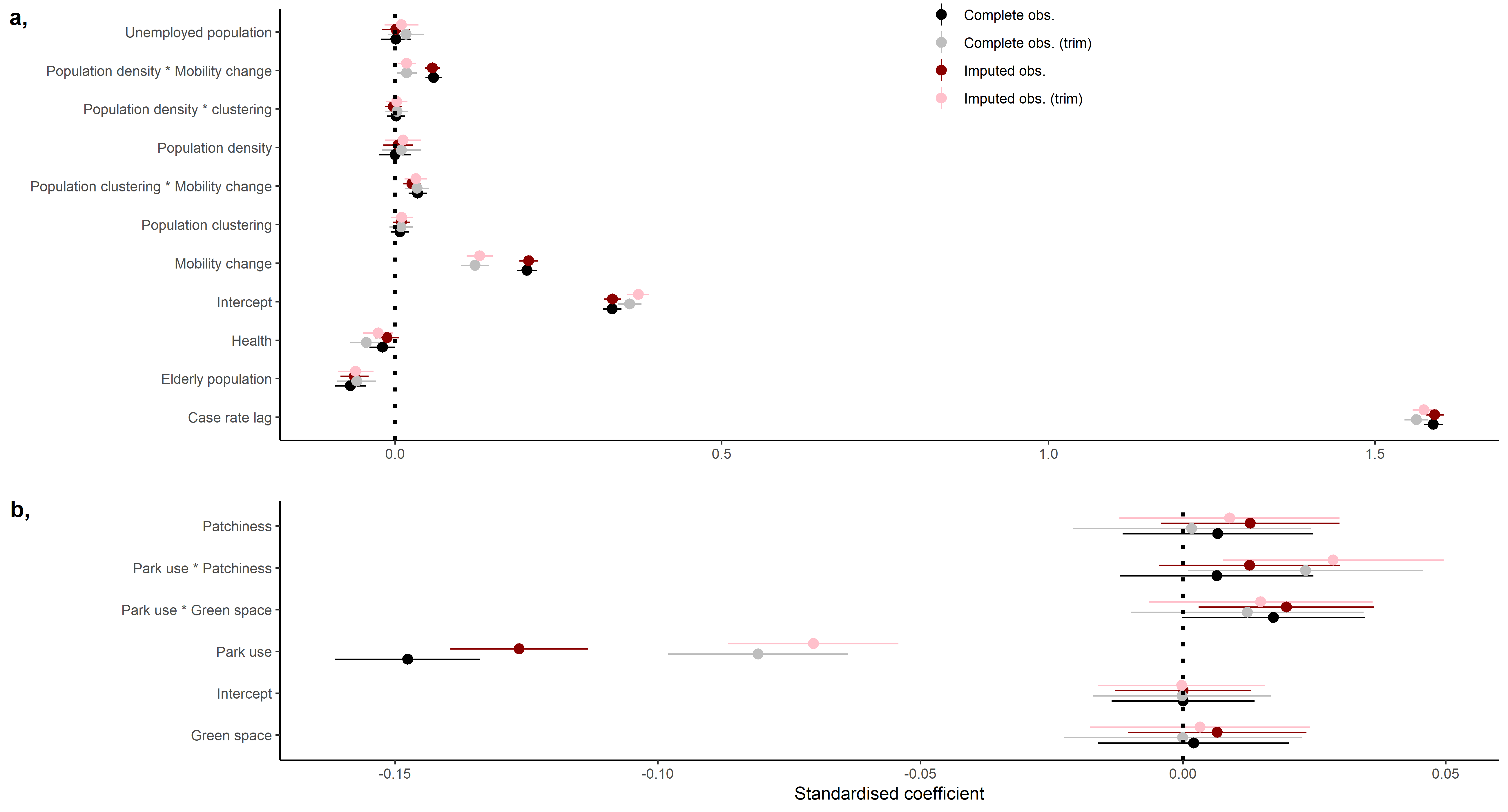

**Figure S5.** Standardised coefficients of the baseline (a) and green (b) transmission models with (monochrome) and without (shades of red) imputed values. The darker shade (black and dark red) of grouping represents the coefficients from the full temporal extent of the dataset (March 1^st^ to November 30^th^ 2020), while the lighter shade represents the trimmed temporal extent (June 1^st^ to November 30^th^ 2020), were the opening months of the pandemic were excluded to reduce the effect of spatial variability in case testing availability. Error bars represent 95% confidence intervals. Note that x-axes extents differ between panels.

**Equation S1**. Baseline transmission model structure with *case rate* as the response *β* representing the model intercept and coefficients. *f* provides random effect smoothing over *local authority* and cyclic smoothing over *day of the week.*

*case rate =*

*β*_0_ *+*

*β_1_*(health) +*

*β_2_*(age) +*

*β_3_*(economy) +*

*β_4_*(population density) +*

*β_5_*(population clustering) +*

*β_6_*(mobility change) +*

*β_7_*(community cases) +*

*β_8_*(population density * population clustering) +*

*β_9_*(population density * mobility change) +*

*β_10_*(population clustering * mobility change) +*

*β_11_*(mobility change * community cases) +*

*f(local authority) +*

*f(day of the week)* +

*ε*

**Equation S2.** Green transmission model structure with *residual case rate* as the response *β* representing the model intercept and coefficients. *f* provides random effect smoothing over *local authority*.

*residual case rate =*

*β*_0_ *+*

*β_1_*(green space) +*

*β_2_*(patchiness) +*

*β_4_*(park use) +*

*β_5_*(green space * park use) +*

*β_6_*(patchiness * park use) +*

*f(local authority) +*

*ε*

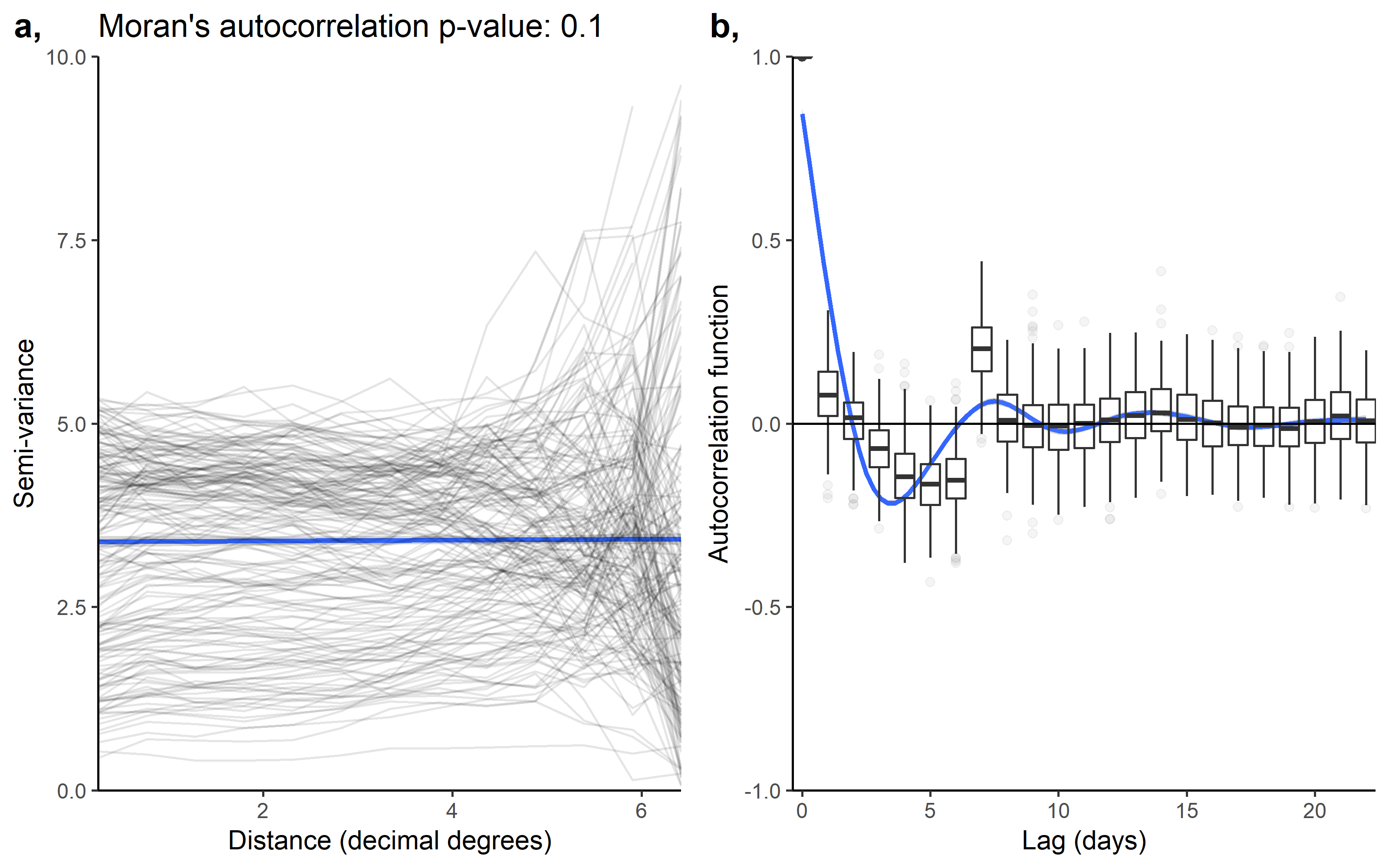

**Figure S6.** a) Spatial autocorrelation of the baseline transmission model residuals, showing the change in semi-variance between local authorities at different distances. Each grey line represents the spatial autocorrelation for different days, whilst the average (locally estimated scatterplot smoothing) spatial correlation is represented by the blue line. We report the median (across days) Moran’s autocorrelation in the plot title. b) Temporal autocorrelation of baseline transmission model, with each boxplot describing the median and interquartile range of temporal autocorrelation across local authorities. The blue line describes the average (locally estimated scatterplot smoothing) temporal autocorrelation over different lags.

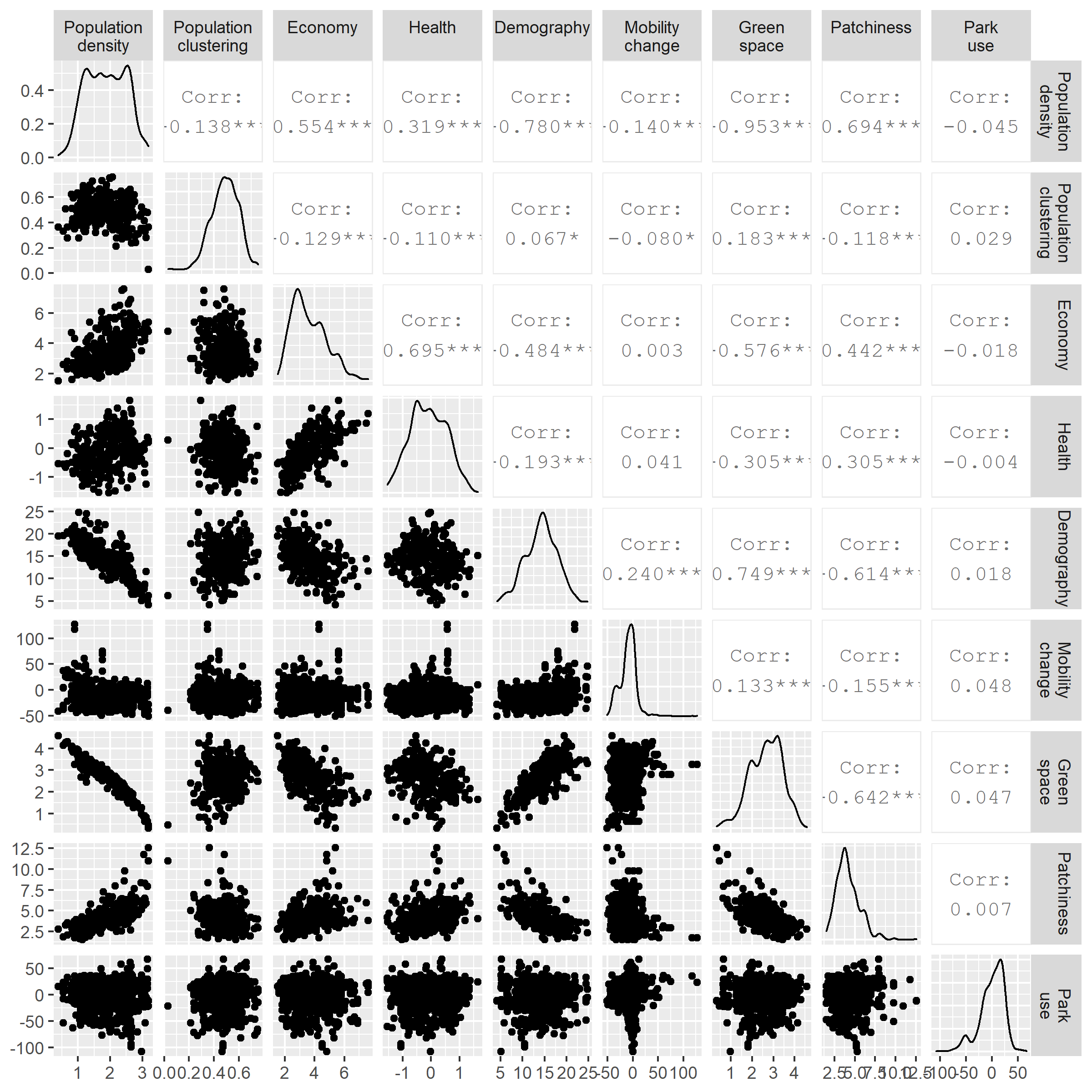

**Figure S7.** Correlation matrix of predictors in the baseline and green transmission models, with scatterplots beneath the diagonal, density plots of the predictor on the diagonal, and Pearson correlation coefficients (where * = significant at p0.05, ** = significant at p = 0.01, and *** - significant at p = 0.001) above the diagonal.
